# Covid-19 clinical data analysis using Ball Mapper

**DOI:** 10.1101/2020.04.10.20061374

**Authors:** Pawel Dlotko, Simon Rudkin

## Abstract

In this note we provide a result of analysis of blood test data from patients with SARS-Cov-2 using Ball Mapper Algorithm. We observe that patients with the virus and in particularly patients who end up in Intensive Care Unit have quite narrow values of those parameters. Please note that this is a preliminary work and it need to be validated on much larger dataset which we are trying to acquire at the moment.

## 1 Introduction

This is a work in progress. We share the results now hoping that they may be useful for people doing similar type of analysis to ours. If you have any questions or comments, please email us. Please note that this paper will be edited, and it is far from its final version

Covid-19 rapidly becoming established as the modern global pandemics of the twenty first century. It is currently causing a large number of deaths, serious social distraction and major economical losses in the world especially in the Western Europe and the USA.

To response this major treat word-wide scientific community is mobilising to use scientific methods to fight this pandemic. In this note we will analyse the clinical data made available by Hospital Israelita Albert Einstein in Sao Paulo, Brasil. The anonymyzed dataset is provided at Data4u (2020).

In the analysis we will use methods of *Topological Data Analysis*. Starting from pioneering works as in Carlsson (2009) and Nicolau et al. (2011) we will build up on the recent developement presented in Dlotko (2019a) to localize clusters of patients that may be important from the perspective of a healthcare provider. We demonstrate that within the measurements of 16 major blood characteristics there is significant information to reliably classify those patients who will require ICU treatment, isolate those spaces where Covid-19 is most active and hence obtain a quick forecast of the likely care requirements of a presenting individual.

## 2 Dataset

The available dataset is composed of the following data collumns:

1. Patient age (integer quantile), ranging between 0 and 19,
2. SARS-Cov-2 exam result – binary variable having values *positive* or *negative*,
3. Patient addmited to regular ward (1=yes, 0=no),
4. Patient addmited to semi-intensive unit (1=yes, 0=no),
5. Patient addmited to intensive care unit (1=yes, 0=no),
6. Hematocrit, ranging between −4.501419544 and 2.662703753,
7. Hemoglobin, ranging between −4.345602989 and 2.671867847,
8. Platelets, ranging between −2.5524261 and 9.53203392,
9. Mean Platelet volume, ranging between −2.457574606 and 3.713052034,
10. Red blood Cells, ranging between −3.970608234 and 3.645706177,
11. Lymphocytes, ranging between −1.865069628 and 3.764099598,
12. Mean corpuscular hemoglobin concentration (MCHC), ranging between −5.431808472 and 3.331070662,
13. Leukocytes, ranging between −2.020302534 and 4.522041798,
14. Basophils, ranging between −1.140143752 and 11.07821941,
15. Mean corpuscular hemoglobin (MCH), ranging between −5.937603951 and 4.098546028,
16. Eosinophils, ranging between −0.835507691 and 8.350875854,
17. Mean corpuscular volume (MCV), ranging between −5.101581097 and 3.410979986,
18. Monocytes, ranging between −2.163721323 and 4.533397198,
19. Red blood cell distribution width (RDW), ranging between −1.598094344 and 6.982183933.

In this work we will use variables from columns 1 as well as 6-20 as a characteristics/descriptor of a patient. Our target will be to locate any clusters of patients with particularly high value of variable 2 (positively of SARS-Cov-2 result), patients that have been admitted to a regular ward, semi intensive or intensive care unit. For the purpose of this study we will use Ball Mapper algorithm Dlotko (2019a) and its public domain implementation Dlotko (2019b) available through CRAN repository. To make the obtained results reproducible, all the code that have been used, will be presented in this paper.

In order to reproduce the results of this paper the Reader should install R language R Core Team (2019) and the BallMapper package Dlotko (2019b). Subsequently the dataset is to be downloaded from the Kaagle webpage Data4u (2020). This dataset is available in the format of Microsoft Exel. For the fuhrer processing it should be stored as a CSV file.

## 3 Ball Mapper and TDA

The Ball Mapper algorithm, inspired by the work in Carlsson (2009), is a tool to produce a topological faithful representation of high dimensional dataset that can be visualized and explored in two dimensions. To achieve this aim, an input collection of points *P* together with a distance between points *d*, is covered with a collection of balls of a radius ∈ > 0. The centers of balls *c*_1_, …, *c*_*n*_ are selected from the points of *P* in a greedy way as described in Dlotko (2019a).

The fact that the whole point cloud *P* is covered by balls of a radius ∈ centered in *c*_1_, …, *c*_*n*_ implies that for every *x* ∈ *P* there exist *i* ∈ {1, …*n*} such that *d*(*x, c*_*i*_) ≤ ∈.

Given such a cover an abstract graph is constructed in the following way: Vertices corresponds to the centers of balls *c*_1_, …, *c*_*n*_. An edge is placed between vertices *c*_*i*_ and *c*_*j*_ for *i, j* ∈ {1, …, *n*} if there exist *p* ∈ *P* such that *d*(*p, c*_*i*_) ≤ ∈ and *d*(*p, c*_*j*_) ≤ ∈. In other words, the point *p* is covered jointly by the ball centered in *c*_*i*_ and the ball centered in *c*_*j*_.

The graph obtained as described above is called a *Ball Mapper graph*. Its layout is a manifestation of the layout of the high dimensional point could *P*. Sizes of vertices in its visualization will correspond to the number of points covered by the corresponding ball.

In this experiment the variables 1 and 6-20 will constitute the point cloud *P* used to create the Ball Mapper graphs analyzed in this paper. A standard Euclidean distance between points will be used.

Our task is to present how the predictive variables 2, 3, 4 and 5 (SARS-Cov-2 result, standard ward, semi intensive care and intensive care admission) changes over *P* by examining how they change over the obtained Ball Mapper graph. This will be achieved by the following procedure: every vertex of the Ball Mapper graph corresponds to a ball *B*(*c*_*i*_, ∈). Let us take *P*_*i*_ = *B*(*c*_*i*_, ∈)∩*P*. The value of the ball, and therefore a point in the Ball Mapper graph, is determined as an average of value of the characteristic (one of the variables 1-5) in *P*_*i*_. This value will be presented using colour scale, where red colours means low values and violet means high values of the predictive variable.

## 4 Raw Data

As one can observe in the Section 2 there is no major variation between ranges of variables except from the first variable. Therefore in this section we will use the original data from Data4u (2020) without any normalization.

In the first step the data are read into an array into R. We will also remove the rows with missing values and convert categorical variable in the column 2 to a numeric one.

~~~
library(‘BallMapper’)
data <- read.csv(‘dataset.csv’, header = TRUE)
data <- data[2:20]
data <- na.omit(data)
data[2] <- 1-as.numeric(data[2] == ‘negative’)
~~~

Now, the data need to be re-arranged into a point cloud *P* and exploratory variables. This is achieved using the following code:

~~~
covid_test <- data[,2]
regular_ward <- data[,3]
semi_intensive_care <- data[,4]
intensive_care <- data[,5]
covid_and_icu <- covid_test*intensive_care
pt_cloud<-cbind(data[,1],data[,6:19])
~~~

Give the prepared datasets, we can create the initial Ball Mapper plots using the following command. The radius ∈ set to 7 was chosen to belance the level of detail in the plot with the need for a tangible discussion of the evidence.

~~~
BM1 <- BallMapper::BallMapper(as.data.frame(pt_cloud),as.data.frame(covid_test),7)
BallMapper::ColorIgraphPlot(BM1,seed_for_plotting = 123)
~~~

As a result the following plot presented in the Figure 1 is obtained. The obtained Ball Mapper graph suggest that the patients, which are likely to have positive result for SARS-Cov2, have quite similar values coming from the blood tests. It is straightforward to give the values of all the used variables for clinical practitioners. As an example, we will consider all the balls 1 and 10 it can be achieved by the following instructions:

**Figure 1:**
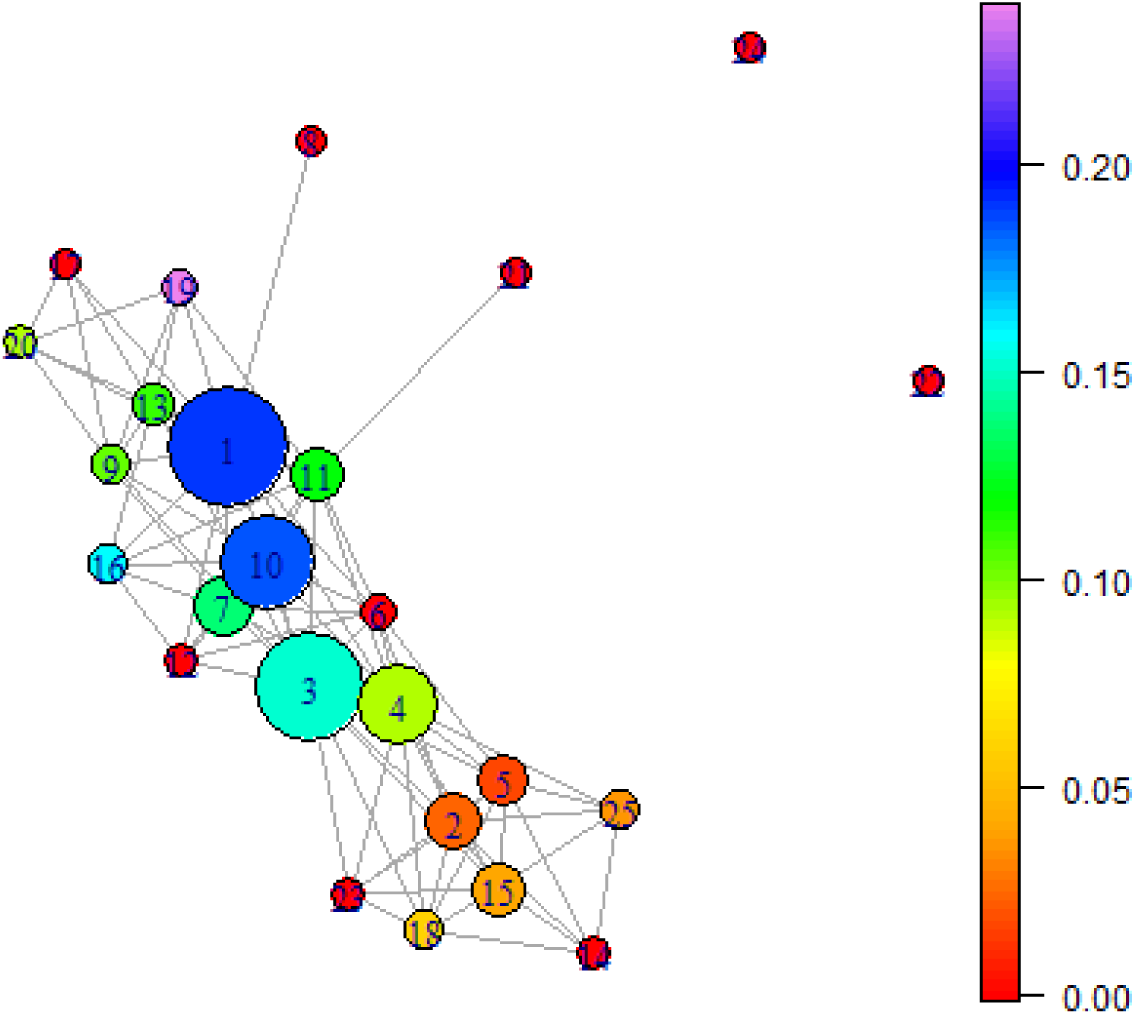
Ball Mapper plot for not normalized dataset with radius 7 colored by the result of Covid-19 test.

~~~
pts <- BallMapper::points_covered_by_landmarks(BM1, c(1,10))
subset <- data[pts,]
av <- numeric(length(subset))
stdev <- numeric(length(subset))
for (i in 1:length(subset))
{
  av[i] <- mean(subset[,i])
  stdev[i] <- sd(subset[,i])
}
~~~

The obtained values of average and standard deviations are summarized in the array below:

| Measurement | av | stdev |
| --- | --- | --- |
| Patient age quantile | 13.92821782 | 3.6158317 |
| SARS-Cov-2 exam result | 0.175742574 | 0.3810729 |
| regular ward | 0.111386139 | 0.3149998 |
| semi-intensive unit | 0.061881188 | 0.2412384 |
| intensive care unit | 0.024752475 | 0.1555625 |
| Hematocrit | 0.206828096 | 0.8017905 |
| Hemoglobin | 0.186720024 | 0.8143198 |
| Platelets | -0.110522314 | 0.82834 |
| Mean platelet volume | 0.073160018 | 0.9683485 |
| Red blood Cells | 0.12583136 | 0.8406223 |
| Lymphocytes | -0.006709115 | 0.9116392 |
| MCHC | -0.02500837 | 0.8880864 |
| Leukocytes | -0.130166666 | 0.8178861 |
| Basophils | 0.034815154 | 0.8037858 |
| MCH | 0.081265496 | 0.7504845 |
| Eosinophils | -0.019005055 | 0.8222541 |
| MCV | 0.107442011 | 0.7604749 |
| Monocytes | 0.05668995 | 0.940488 |
| RDW | -0.081415821 | 0.7258069 |

In out second experiment we will use the value of the column 5 (determining if a patient was admitted to an intensive care unit). For that purpose the following code is used:

~~~
BM2 <- BallMapper::BallMapper(as.data.frame(pt_cloud),as.data.frame(intensive_care),7)
BallMapper::ColorIgraphPlot(BM2,seed_for_plotting = 123)
~~~

The results are presented in the Figure 2.

**Figure 2:**
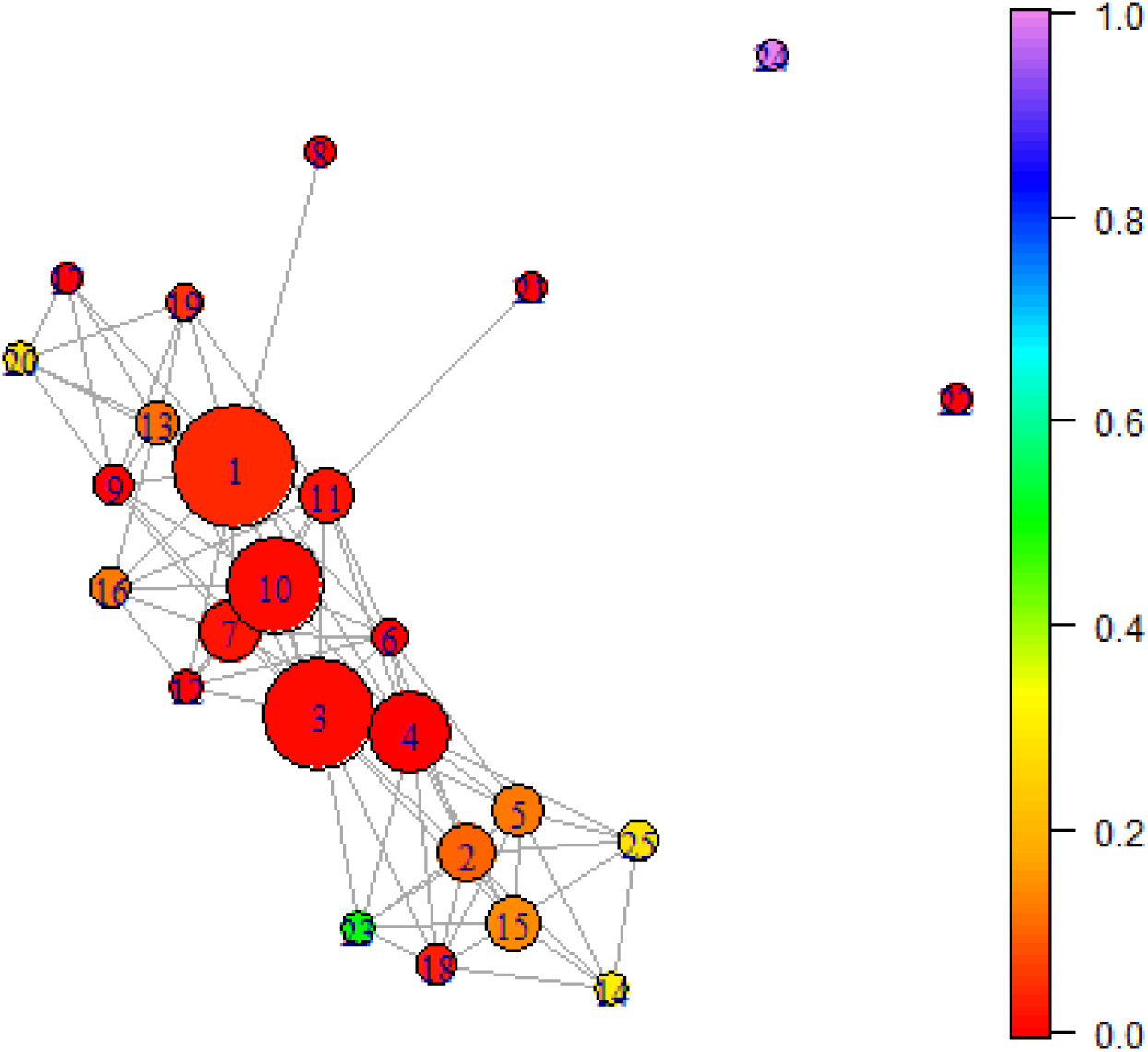
Ball Mapper plot for not normalized dataset with radius 7 colored by the variable determined if the patient was admitted to a Intensive Care Unit (ICU).

As one can observe, the patients in the balls 14,23,24,24 but also 20 and 13 are likely to end up in the ICU. It is worth noticing that those balls are far away in space, and therefore they represent patients with different values of blood parameters that are used to construct the Ball Mapper graph. Once gain, it is possible to recover the precise values of be blood tests parameters and use them to assess the risk for newly admitted patients.

Let us not consider the patients which are tested positively for SARS-Cov-2 that required treatment in Intensive Care Unit. To visualize their location the following code is to be used:

~~~
BM3 <- BallMapper::BallMapper(as.data.frame(pt_cloud),as.data.frame(covid_and_icu),7)
BallMapper::ColorIgraphPlot(BM3,seed_for_plotting = 123)
~~~

As a result the Ball Mapper plot presented in Figure 3 is obtained. As one can observe, Ball number 1 contains a lot of such patients, but also ball number 16 and 25 is clearly visible and majority of patients over there wil require ICU.

**Figure 3:**
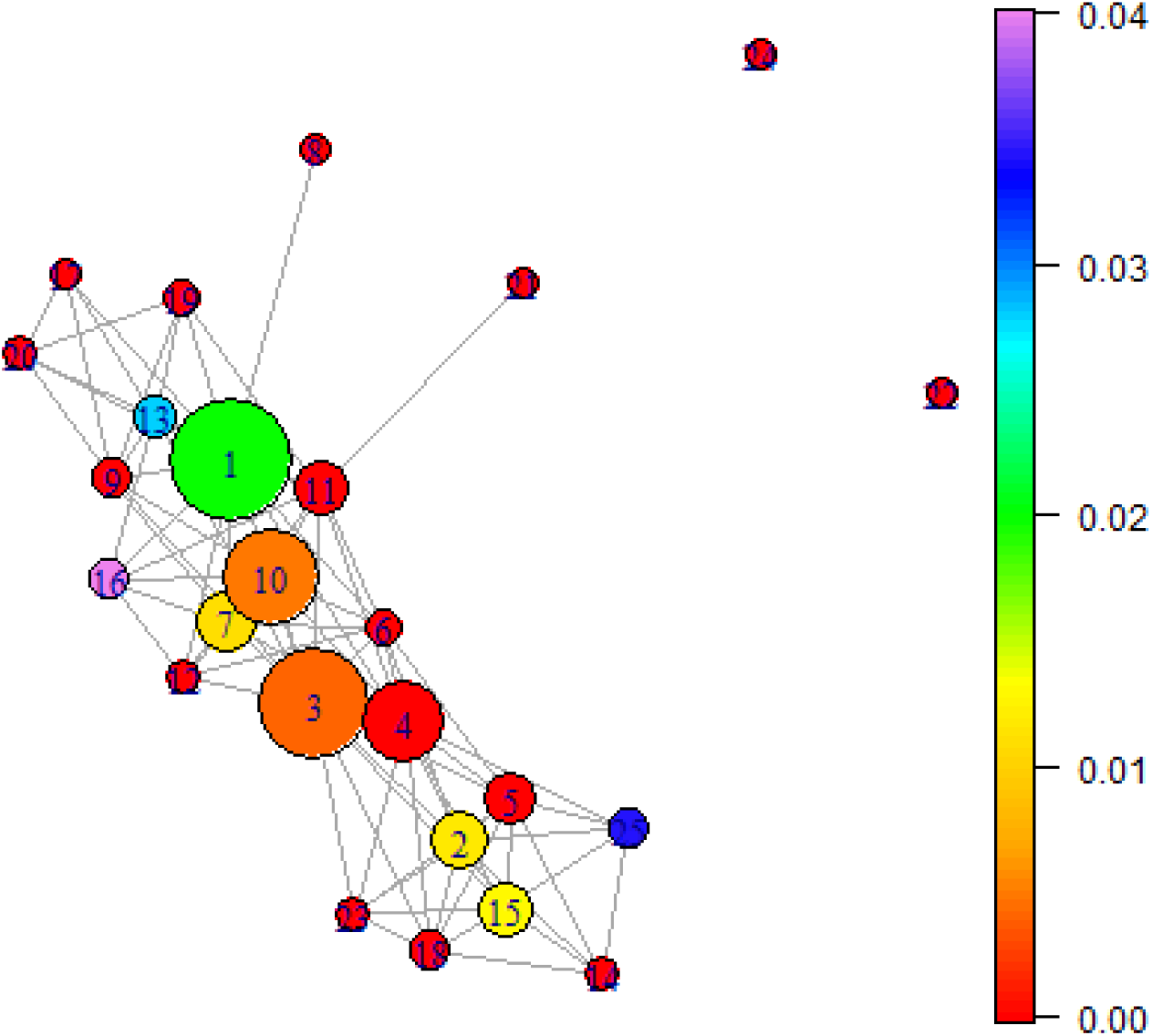
Ball Mapper plot for not normalized dataset with radius 7. Colouration by the patientest that are tested positively for SARS-Cov-2 that required Intensive Care treatment.

In our next exercise we will focus on the patients which have positive values of tests for SARS-Cov2. To obtain them the following code is used:

~~~
covid_positive_data <- data[data[,2]==1,]
covid_positive_regular_ward <- covid_positive_data[,3]
covid_positive_semi_intensive_care <- covid_positive_data [,4]
covid_positive_intensive_care <- covid_positive_data [,5]
covid_positive_pt_cloud<-cbind(covid_positive_data [,1], covid_positive_data[,6:19])
~~~

Let us observe which of those patients end up in normal ward, semi intensive and intensive care. For that purpose the following code is used. Once again, the value ∈ = 5 is chosen to optimise the information tractability trade off.

~~~
BM3 <- BallMapper::BallMapper(as.data.frame(covid_positive_pt_cloud),
as.data.frame(covid_positive_regular_ward),5)
BallMapper::ColorIgraphPlot(BM3,seed_for_plotting = 123)

BM4 <- BallMapper::BallMapper(as.data.frame(covid_positive_pt_cloud),
as.data.frame(covid_positive_semi_intensive_care),5)
BallMapper::ColorIgraphPlot(BM4,seed_for_plotting = 123)

BM5 <- BallMapper::BallMapper(as.data.frame(covid_positive_pt_cloud),
as.data.frame(covid_positive_intensive_care),5)
BallMapper::ColorIgraphPlot(BM5,seed_for_plotting = 123)
~~~

The results are presented in the Figure 4

**Figure 4:**
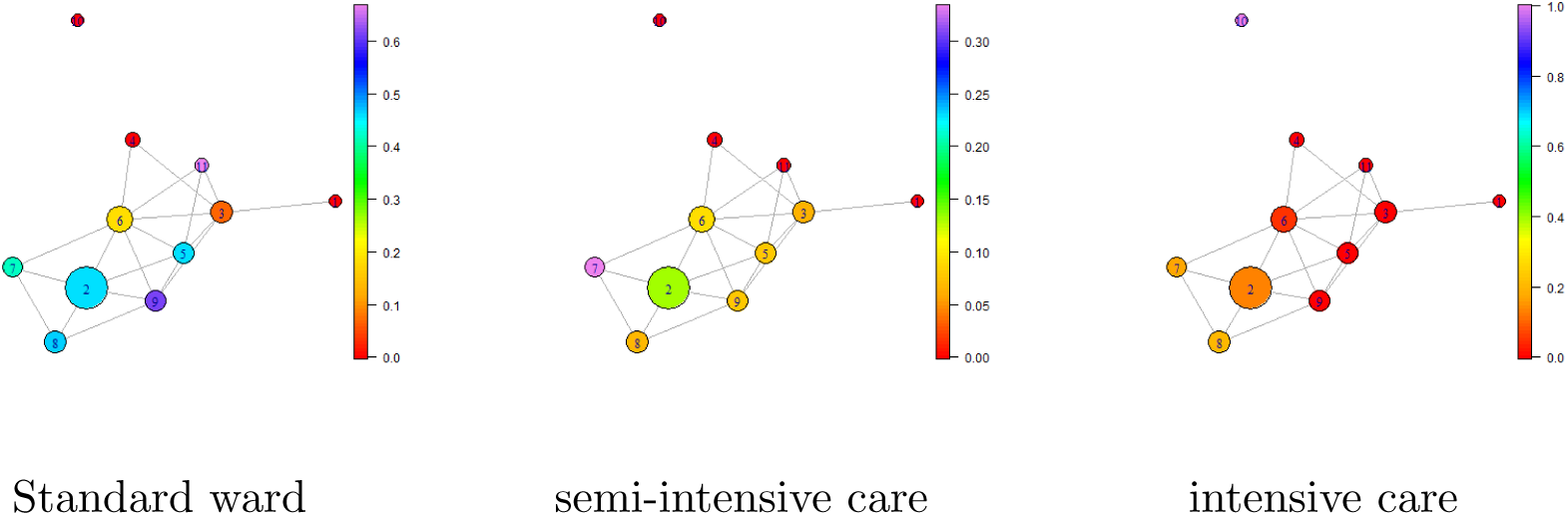
caption

As one can observe, the patients in the Ball 10 almost entirely end up in the intensive care unit as well as 20% of patients from balls 2, 7, 8. Patients from balls 1, 4, 11 will need at most semi intensive care.

In the second version of the paper, guided by a medical expert, we will provide combinations of variables that is characteristic for patients with SARS-Cov-2 as well as the characteristics of patients that will need an intensive care unit.

Unfortunately the dataset do not provide the information about the patients who died (except that there are such patients in the dataset). Consequently we are not able to locate them in the global picture and we cannot assess the risk of death of a patient. Consequently we cannot determine if there is a combination of blood results that is characteristics for those patients.

## 5 Normalized data

In this section we will revisit the experiments from the Section 4 with all the columns normalized to the interval [0, 1]. This will be performed by constructing a linear map that transform the minimal value in the column to 0 and maximal value in the column to 1. This normalization may be important for the following reason: as one can see by the ranges of variables described in Section 2 it may be the case that not normalized data are skewed towards more senior patients as the value of the first columns is, for them, considerably larger than the values of the other columns.

To normalize the data the following code is to be used:

~~~
pt_cloud_normalized <- BallMapper::normalize_to_min_0_max_1(data)
~~~

Given the normalized data we can now see where the SARS-Cov-2 patients as well as the patients that will require a intensive care unit are located in this normalized dataset. Note that, as usual, the value of the parameter ∈ is set to 1.2 in this instance.

~~~
BM6 <- BallMapper::BallMapper(as.data.frame(pt_cloud_normalized),
as.data.frame(covid_test),1.2)
BallMapper::ColorIgraphPlot(BM6,seed_for_plotting = 123)
BM7 <- BallMapper::BallMapper(as.data.frame(pt_cloud_normalized),
as.data.frame(intensive_care),1.2)
BallMapper::ColorIgraphPlot(BM7,seed_for_plotting = 123)
~~~

In the Figure 5 we can see quite different picture compared to the one presented in Figure 1. This time the patients which are SARS-Cov-2 positive values are all located in balls 5,6, 10, 12, 18, while ball 10 contain only patients with positive values of tests. It is worth remarking that the patients with the positive values of tests are located close to a large ball number 1 that contain no patients with SARS-Cov-2.

**Figure 5:**
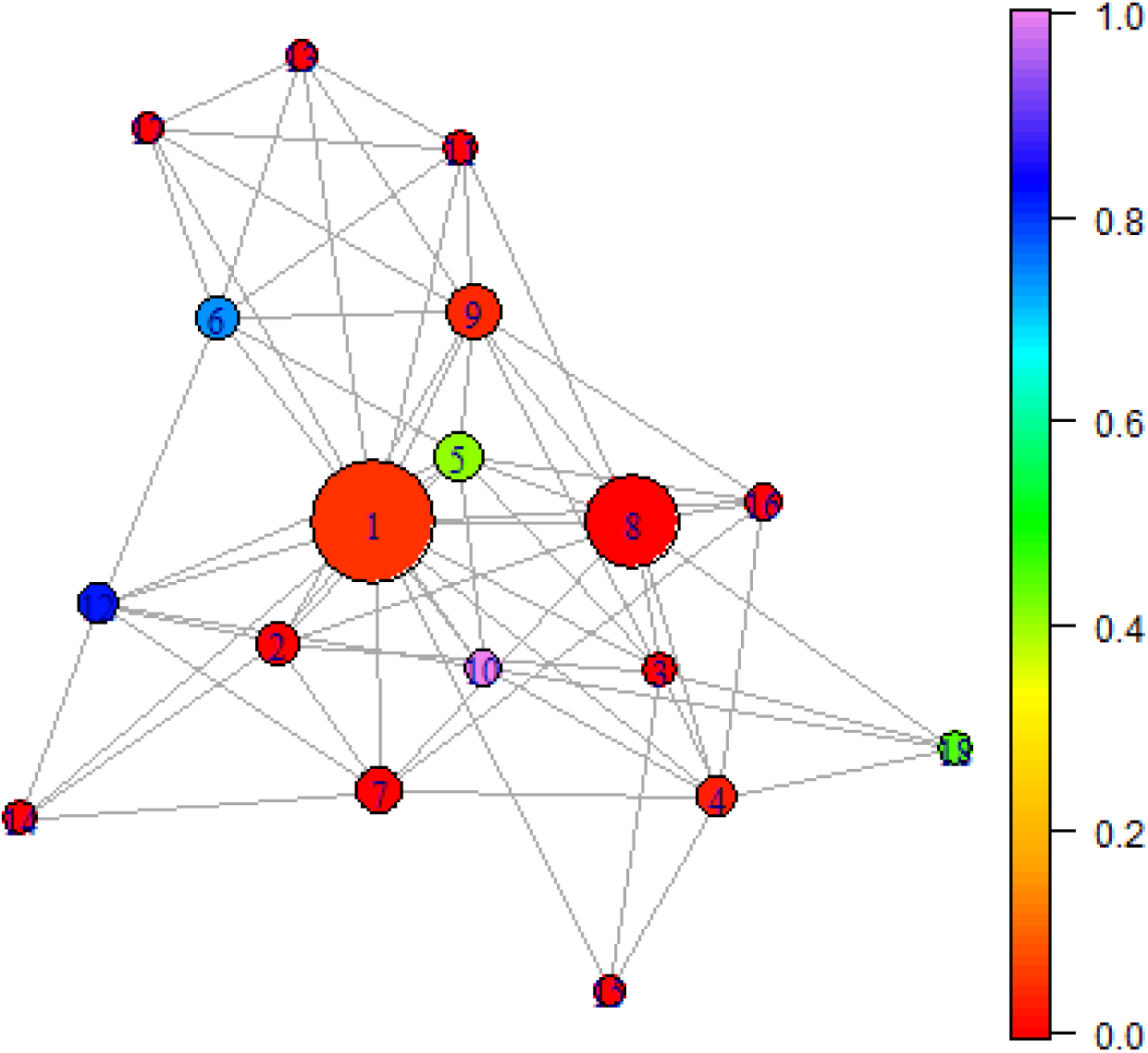
Ball Mapper graph for normalized data colored by the result of test for SARS-Cov-2.

This observation may indicate some relation between blood results and SARS-Cov-2 that may be useful to locate high risk groups and speed up diagnosis using only standard blood tests.

Figure 6 show which patients required an Intensive Care Unit (ICU). As one can observe, amount the patients tested positively for SARS-Cov-2 the patients in the balls 10 and 18 have required ICU, but patients from other balls did not. Please note that the patients in Balls 3, 4 and 15 also required ICU, however those balls did not contain majority of SARS-Cov-2 patients. We believe that this is because the data describe all the patients in the hospital, not only ones with SARS-Cov-2.

**Figure 6:**
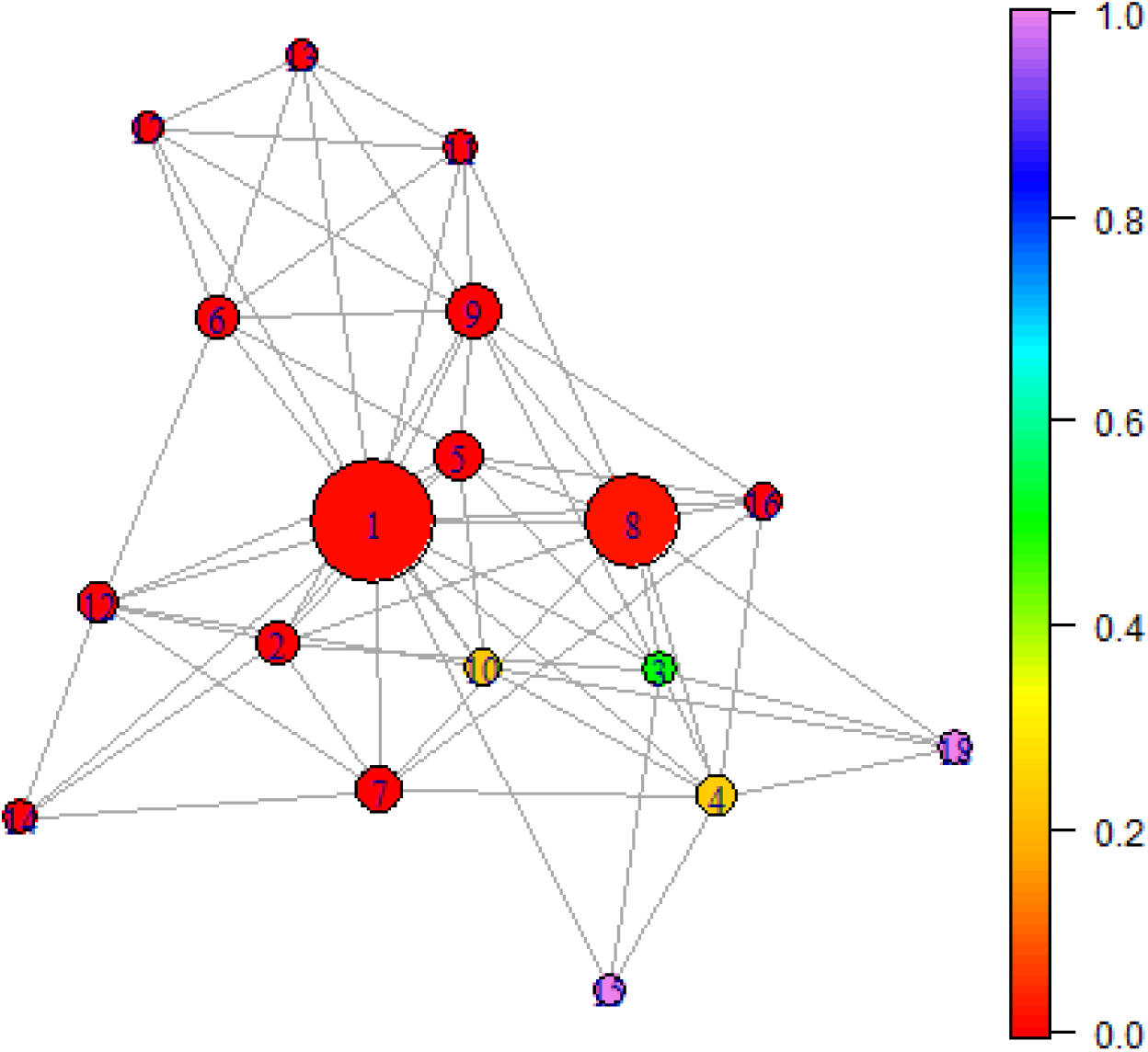
Ball Mapper graph for normalized data colored by the patients who required Intensive Care Unit.

To locate the patients with SARS-Cov-2 that were admitted to ICU the following code can be used:

~~~
BM6 <- BallMapper::BallMapper(as.data.frame(pt_cloud_normalized),
as.data.frame(covid_and_icu),1.2)
BallMapper::ColorIgraphPlot(BM6,seed_for_plotting = 123)
~~~

The results can be found in the Figure 7. Once again, we see clear patterns in balls 10 and 18 of patients tested positively for SARS-Cov-2 that require Intensive Care treatment. In the medical practice, they need to be given special attention.

**Figure 7:**
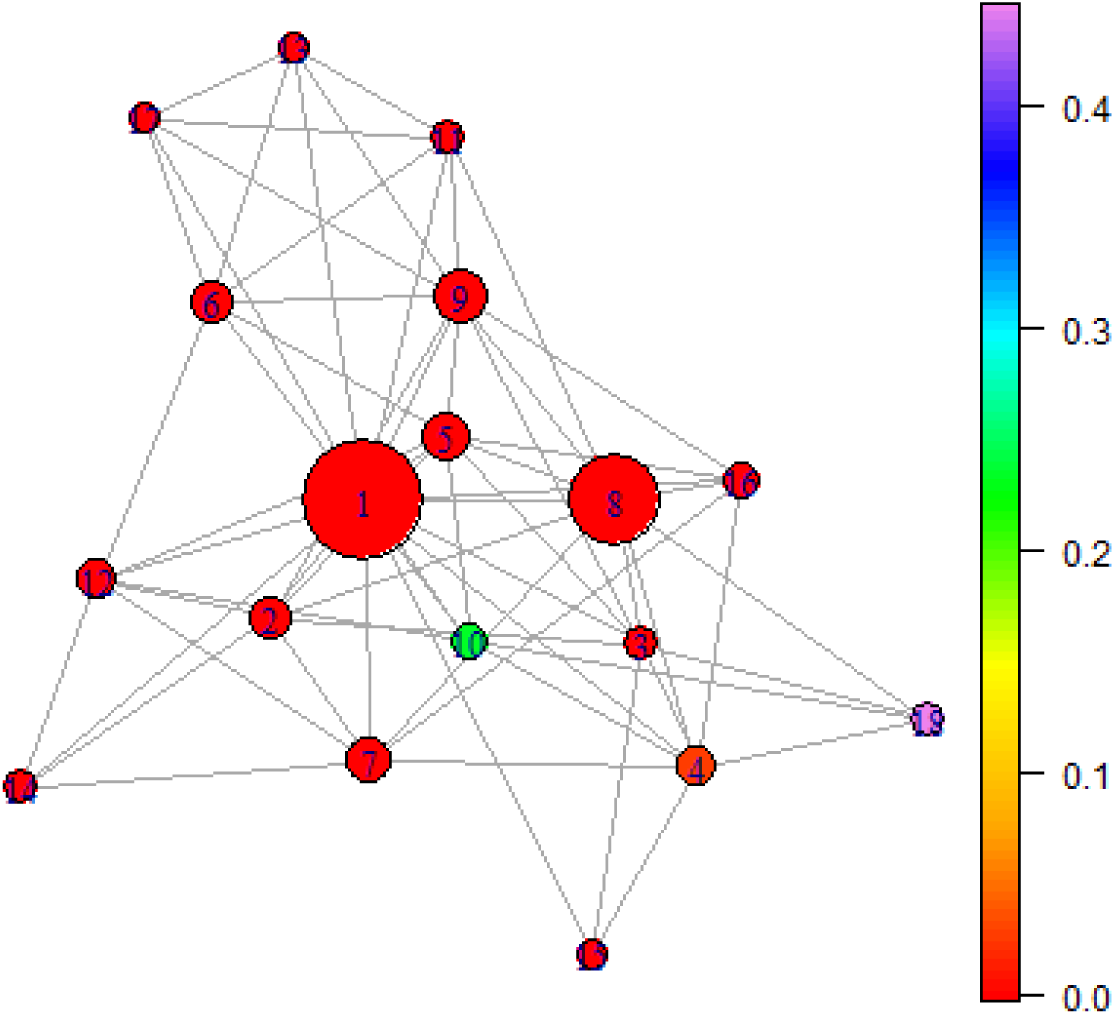
Ball Mapper graph for normalized data colored by the patients who are tested positively for SARS-Cov-2 and required Intensive Care Unit treatement.

In the final part of this analysis we will therefore restrict only to patients diagnosed with SARS-Cov-2. To obtain them the following code is to be used:

~~~
normalized_covid_positive_data <- pt_cloud_normalized[pt_cloud_normalized $ V2==1,]
normalized_covid_positive_regular_ward <- normalized_covid_positive_data [,3]
normalized_covid_positive_semi_intensive_care <- normalized_covid_positive_data [,4]
normalized_covid_positive_intensive_care <- normalized_covid_positive_data [,5]
normalized_covid_positive_pt_cloud<-cbind(normalized_covid_positive_data[,1],
normalized_covid_positive_data[,6:19])
~~~

Given those datasets let us locate the patients in regular ward, Semi Intensive and Intensive Care Units. They can be localized using the following code. As usual, the value ∈ = 1.1 was obtained for the best balance of information and readability.

~~~
BM8 <- BallMapper::BallMapper(as.data.frame(normalized_covid_positive_data),
as.data.frame(normalized_covid_positive_regular_ward),1.1)
BallMapper::ColorIgraphPlot(BM8,seed_for_plotting = 123)

BM9 <- BallMapper::BallMapper(as.data.frame(normalized_covid_positive_data),
as.data.frame(normalized_covid_positive_semi_intensive_care),1.1)
BallMapper::ColorIgraphPlot(BM9,seed_for_plotting = 123)

BM10 <- BallMapper::BallMapper(as.data.frame(normalized_covid_positive_data),
as.data.frame(normalized_covid_positive_intensive_care),1.1)
BallMapper::ColorIgraphPlot(BM10,seed_for_plotting = 123)
~~~

The results are presented in the Figure 8

**Figure 8:**
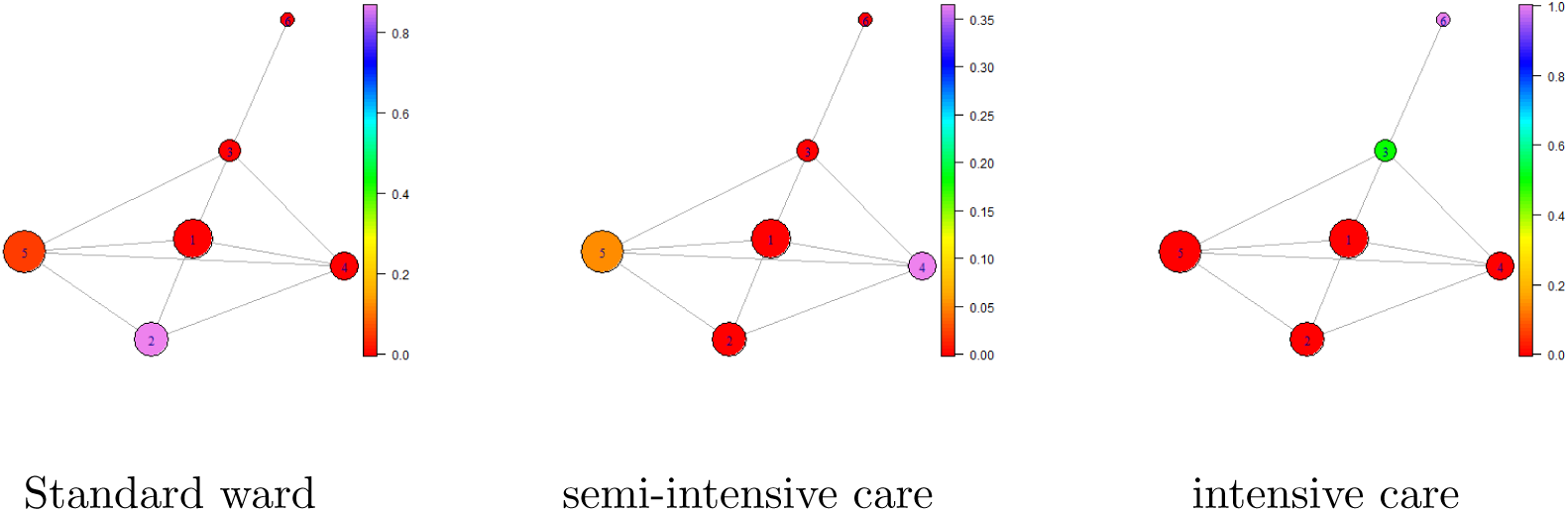
caption

The analysis in the Figure 8 indicate a hierarchy of patients:

1. Those within Ball 2 are very likely to stay in the regular ward.
2. Patients in ball 4 and 5 will probably need semi-intensive care.
3. Patients in balls 3 and 6, especially in 6, will need full ICU.

The presented analysis of the normalized data indicate that blood–based bio-markers may allow to determine, at early stage, patients with higher risk of being tested positively for SARS-Cov-2 as well as those, who may require ICU.

## 6 Conclusions

In the presented note we have analyzed the data provided in Data4u (2020) using methods of Topological Data Analysis, most notably Ball Mapper.

The presented visualization indicate that the patients that may require special care (those with SARS-Cov-2 as well as those who may require ICU) are located in distinct regions of space. Given this, it seems to be possible to build a predictive system that would allow, for a given patient admitted to the hospital, to put him into appropriate risk factor groups. That may allow the health care providers more efficient use of resources and help the patient to optymize the treatment.

## Data Availability

Data are available via Kagle link provided in the paper.

